# Depressed Proximal Glycolysis in Myocardium Of Human Heart Failure with Preserved Ejection Fraction

**DOI:** 10.1101/2023.09.30.23296261

**Authors:** Navid Koleini, Mariam Meddeb, Mohammad Keykhaei, Seoyoung Kwon, Liang Zhao, Virginia Hahn, Kavita Sharma, Erika L Pearce, David A Kass

## Abstract

Heart failure with preserved ejection fraction (HFpEF) accounts for >50% of all heart failure world-wide and remains a major unmet medical need. The most effective recently approved treatments were first developed for diabetes, suggesting metabolic defects are paramount. Myocardial metabolomics in human HFpEF has identified reduced fatty acid and branched chain amino acid catabolism, but the status of glycolysis is unknown. Here we performed targeted metabolomics and protein analysis of glycolytic pathway enzymes in myocardial biopsies of patients with HFpEF versus HF with reduced ejection fraction (HFrEF0 or non-failing controls. Glucose was increased in HFpEF myocardium, but immediate downstream glycolytic metabolites (glucose-6 phosphate, fructose 1,6 diphosphate), were more reduced in HFpEF than the other groups, as were their associated synthetic enzymes hexokinase and phosphofructokinase. Pyruvate was also reduced in HFpEF versus controls. These changes were either not present or substantially less so in HFrEF. Suppression of proximal glycolysis was also coupled to lower metabolites and proteins in the pentose phosphate pathway but was independent of diabetes or obesity. These findings support marked metabolic inflexibility in HFpEF and identifies very proximal blockade in glucose metabolism. Efforts to improve metabolic use of carbohydrates in HFpEF will likely need to target these proximal glycolytic enzymes.

---

Heart failure with preserved ejection fraction (HFpEF) reflects half of all heart failure and remains a major unmet medical need given few effective treatments. It involves multiple organs, and its current primary co-morbidity is obesity/metabolic syndrome^1^. Intriguingly, the most effective approved therapies to date were developed for diabetes^2^ highlighting the importance of understanding metabolic abnormalities underlying HFpEF.

Metabolomic analysis of HFpEF endo-myocardium found depressed fatty-acid oxidation as in HF and a reduced EF (HFrEF)^3^. This was surprising as the HFpEF patients were very obese, a condition thought to augment myocardial fat catabolism. Glycolysis was not assessed; but is generally increased in animal models of hypertrophy, obesity, and HF^4^, and is traditionally viewed as upregulated in HFrEF to offset lower fat catabolism^4^. Prior HFrEF myocardial studies found increased glucose but glycolysis intermediates and related enzymes were not generally altered^5^. To our knowledge, there are no prior data on myocardial glycolytic intermediates and related proteins in human HFpEF.

We performed glycolysis-focused metabolomics of RV-septal endomyocardial biopsies from human subjects with HFpEF (n=24, 63% with diabetes), HFrEF (n=19, 26% with diabetes), or non-failing controls (NF, n=13). Protein expression of pathway members was assessed by immunoblot (normalized to total protein; n=5 NF, n=6 each HF group). Statistical analysis performed with GraphPad Prism Ver 10. Clinical features of each group have been reported^3^. The biopsy protocol was approved by the JHU Institutional Review Board, and all participants provided informed consent.

**Figure panel A** shows the glycolysis pathway and associated pentose phosphate pathway (PPP), with metabolites and protein expression exhibiting significant changes denoted by colored arrows. HFpEF myocardium had greater glucose and expression of membrane glucose transporters Glut-1 and Glut-4 (**B,C**) versus control or HFrEF. Thereafter, however, glycolytic intermediates were significantly lower in HFpEF. The initial step forms glucose 6-phosphate (G-6P) trapping glucose within the cell. G-6P and its generating hexokinases HK1 and HK2 were all significantly reduced in HFpEF (**B,C**). While HKs were unchanged in HFrEF, G-6P was reduced though less than in HFpEF.

**Figure:**
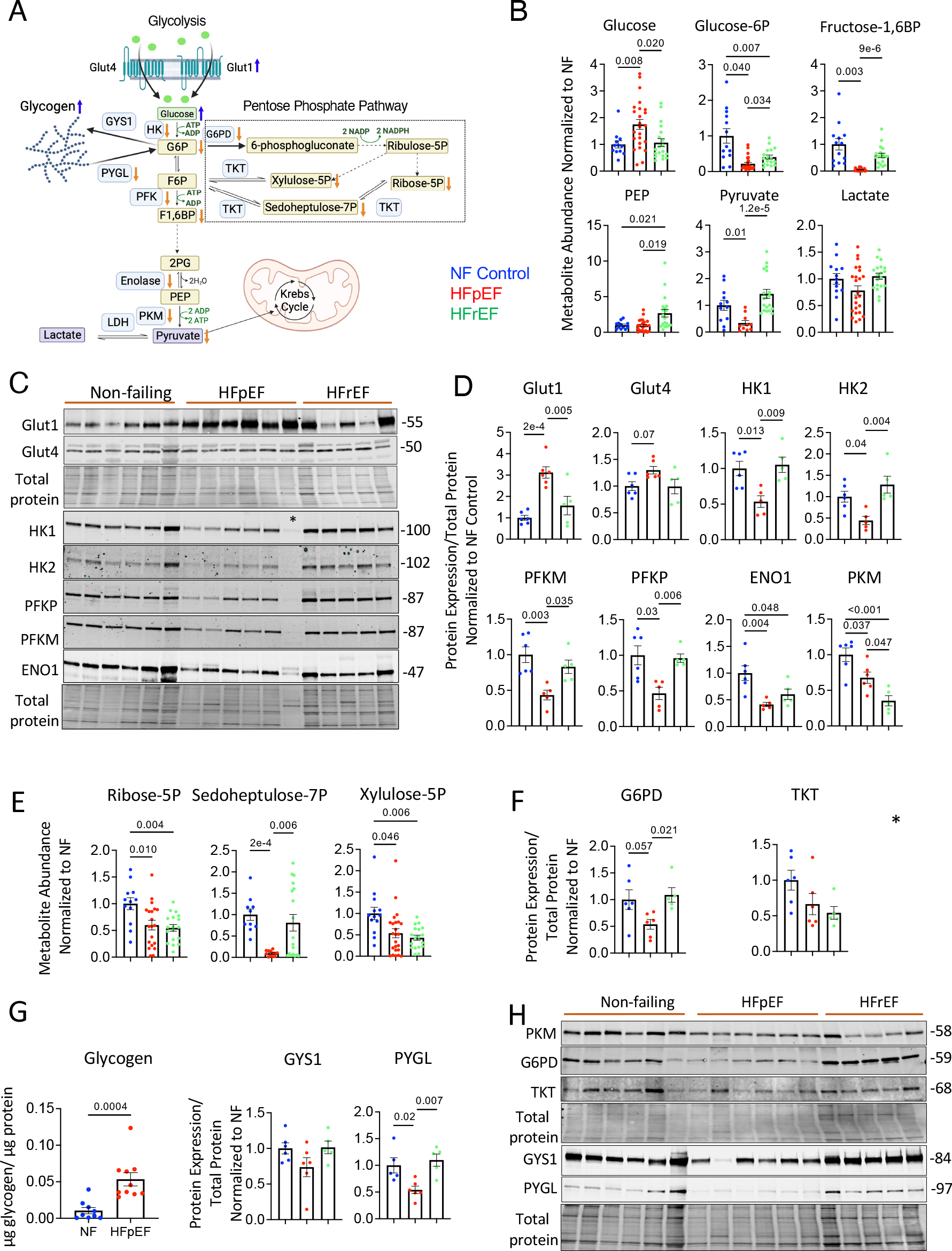
**A)** Schematic diagram of glycolysis and pentose phosphate pathway (PPP) showing results for various metabolic intermediates determined by mass spectrometry and their associated proteins measured by Western Blot. Metabolites or proteins with reduced levels are shown with orange arrows, those increased with blue arrows. LDH – lactate dehydrogenase; 2PG – 2-phosphoglyceric acid; F-6p – fructose 6 phosphate, all other abbreviations as defined in the text. **B)** Summary results for myocardial metabolites in the three patient groups. Data analyzed with 1-way ANOVA or Forsyth-Welch ANOVA if variances differed between groups; P values are provided for multiple comparisons testing using Dunnet’s test. **C)** Immunoblots for glycolytic protein expression, with the corresponding total protein (used for normalization) for each respective gel shown below them. Lane 12 (denoted with *) failed to load and was not used for analysis. **D)** Corresponding summary densitometry of these immunoblots; protein expression normalized to total protein, and then to the result for the non-failing control group. Summary statistics are as described for panel B. **E)** Summary data for PPP metabolites; statistics as described in panel B. **F)** Summary immunoblot densitometry for proteins involved with PPP based on Western blots shown in panel **H**. * P=0.17 between NF and HFpEF, P=0.1 between NF and HFrEF by Kruskal Wallis test and Dunn’s multiple comparisons test. **G)** Modulation of myocardial glycogen. *Left*: total glycogen content in myocardium of NF (n=9) and HFpEF (n=10) hearts, P value Mann Whitney test; *Center, Right:* Summary of protein expression for glycogen synthase 1 (GYS1) and glycogen phosphorylase (PYGL); statistics as in panel B. **H)** Western blot for pyruvate kinase, and proteins in PPP and glycogen pathway. Antibodies from Proteintech were used for Glut1 (21829-1-AP), Glut4 (66846-1-Ig), HK1 (19662-1-AP), PFKM (55028-1-AP), PFKP (68129-1-Ig), ENO1 (11204-1-AP), PKM (10078-2-AP), G6PD (25413-1-AP), TKT (11039-1-AP), GYS1 (10566-1-AP), and PYGL (15851-1-AP). HK2 antibody was from Cell Signaling (2867S).

The subsequent rate limiting step in glycolysis is formation of fructose 1,6 biphosphate (F-1,6-BP) by phosphofructokinases (PFKM, PFKP). Both kinases were lower in HFpEF but not HFrEF (**B,C**) and F-1,6-BP markedly reduced in HFpEF (**B**). Glycolysis ultimately generates pyruvate which can be used for oxidation (ATP synthesis) or generation of lactate. Its immediate precursor is phosphoenolpyruvate (PEP) which we found increased only in HFrEF (**B**). However, expression of enolase-1 (ENO1) and pyruvate kinase (PKM) converting PEP to pyruvate were significantly lower in both HF groups (**C,D,H**), the latter more in HFrEF. Perhaps as a result, pyruvate was reduced in HFpEF but not HFrEF (**B,C**). Lactate was similar among groups, so depressed pyruvate in HFpEF was unlikely from greater lactate conversion (**B**).

Proximal glycolytic metabolic intermediates can be shunted into alterative pathways such as the PPP to provide alternative metabolites and a means to circumvent glycolytic block. This is fueled by key proximal metabolites and specific enzymes. We found PPP intermediates xylulose-5P, ribose-5P, and sedoheptulose-7P significantly reduced in HFpEF (**E**), consistent with less expression of Glucose-6P dehydrogenase (**F,H**) and pathway input metabolites. Another PPP enzyme, transketolase (TKT) trended downward with HF, but this did not reach statistical significance.

As glucose was elevated but glycolysis markedly depressed in HFpEF, we measured glycogen and found it higher in HFpEF. Expression of glycogen synthase (GYS1) was not significantly changed, but glycogen phosphorylase (PYGL) converting glycogen to G6P was downregulated in HFpEF (**G**), supporting a shift favoring glycogen storage versus synthesis.

These data reveal profound reduction of proximal glycolytic intermediates and associated metabolism proteins in human HFpEF myocardium that differ considerably from that found in HFrEF. While HFpEF patients were more obese and had more diabetes than HFrEF, none of the metabolites measured correlated with hemoglobin A1C, nor were there mean differences in their levels +/- a history of diabetes. The reduced expression of proteins in glycolysis was broadly concordant with their mRNA expression we reported from similar tissues^1^. The HFrEF vs NF metabolic and protein findings are also generally consistent with a 2022 study using similar tissues^5^. The proximal nature of the glycolytic blockade associated with reduced HK and PFK means that alternative pathways such as the PPP are also compromised, further limiting fuel flexibility. The increase in glycogen is intriguing, and might further contribute to HFpEF disease, although this remains further examination. One caveat to our steady state metabolomic analysis is that levels do not necessarily translate to reduced flux through the pathway. However, finding associated proteins downregulated upstream of each metabolite supports this interpretation. In summary, the current results identify a striking disparity depression of myocardial glycolysis in HFpEF unlike HFrEF, adding further evidence that HFpEF likely has substantial metabolic insufficiency and inflexibility. Any efforts to stimulate glucose utilization in HFpEF will need to offset this proximal blockade.

## Data Availability

Data available from corresponding author upon reasonable request.

## Acknowledgments

Metabolomics were done at Johns Hopkins University, and at Complete Omics Inc, Baltimore, MD. The work is supported by NIH R35:135827, R35:166565, The Belfer Endowment (DAK), American Heart Association Fellowship 23POST1026402 (NK), Amgen Inc. (KS), K23HL166770 (VH), NIAID: R01AI156274 (EP,DAK).

